# Contraceptive efficacy of 50 mg Mifepristone once-a-week: an open label, single-arm, multicenter clinical trial in Moldova

**DOI:** 10.64898/2026.10.27.26366578

**Authors:** Caroline Frisendahl, Agnes Herré, Karin Brandell, Kristina Gemzell Danielsson

## Abstract

**Introduction:** Unmet needs for contraception persist partly due to side effects, dissatisfaction, and medical contraindications. Mechanistic and clinical data support the contraceptive potential of mifepristone, which may provide a non-estrogen contraceptive option, but evidence remains limited. The primary aim of this trial was to evaluate the contraceptive efficacy and safety of once-weekly 50 mg mifepristone.

**Method:** We conducted an open-label, single-arm, multicenter trial in Moldova. Women aged 18-35 years used mifepristone as their sole contraceptive method for 13 cycles. The primary endpoint was pregnancy due to method failure, expressed as a Pearl Index (PI).

**Results:** Of 170 women enrolled between August 2023 and May 2025, 146 initiated treatment. Two pregnancies occurred during 1,187 evaluable cycles, with a perfect-use at-risk PI of 1.55 (95% CI, 0.04–8.64). Increased endometrial thickness occurred in 8.9% (11/123) of participants and progesterone receptor modulator-associated endometrial changes in 10.6% (13/123), which normalized at follow up. No hyperplasia or cellular atypia was detected in any of the endometrial biopsies. Bleeding averaged one day per cycle, and 60% reported no bleeding or spotting at treatment end. Overall, 95.6% were satisfied or very satisfied with the method.

**Conclusions:** Due to early discontinuation of the trial the study did not achieve the planned sample size. The subsequent imprecise estimated PI did not meet the prespecified efficacy criterion. However, the favorable safety profile, bleeding pattern, and high acceptability supports continued development of mifepristone as a contraceptive.

**Key Message:** Evidence for mifepristone as regular contraception remains limited. 50 mg once weekly showed high acceptability, reduced bleeding, and reversible endometrial changes, expanding evidence for its contraceptive development. The contraceptive efficacy of mifepristone requires confirmation in larger trials.

## INTRODUCTION

Unmet needs in family planning persist, and discontinuation or non-use of contraception due to dissatisfaction, side effects, and medical contraindication is common.^1^ Accordingly, innovation of new contraceptive methods to address diverse family planning needs is essential to further reduce maternal mortality rates and to strengthen sexual and reproductive health, rights, and autonomy worldwide.

The progesterone antagonist mifepristone has in combination with a prostaglandin analogue been used for medical abortion since the late 1980s.^2^ Since then, research has emerged supporting the contraceptive effects of mifepristone.^3,4^ Mifepristone acts primarily as an anti-progestin, preventing pregnancy by inhibiting follicular development,^5,6^ ovulation,^7,8^ and endometrial development,^9,10^ depending on the dose and timing of administration.

Mifepristone is unlikely to cause side effects associated with combined (progestin and estrogen) or progestin-only hormonal contraceptives, such as mood disturbances, decreased libido, acne, and weight gain,^3^ and avoids estrogen-related thromboembolic risk. Potential additional benefits include reduced menstrual bleeding, dysmenorrhea, and fibroid-related symptoms, and possible protection against breast cancer.^11–13^ Mifepristone-based contraception may therefore expand contraceptive options and improve acceptability, particularly for individuals with contraindications to or fear of existing hormonal methods.

The contraceptive efficacy of mifepristone has been evaluated in clinical trials, but evidence remains limited and is primarily based on small pilot studies.^10,14–20^ Daily and once-weekly regimens, particularly at higher doses, appear most promising.^16,19,20^ In a study comparing once-weekly doses, 50 mg resulted in fewer bleeding days and more amenorrhea than 25 mg.^20^ Larger high-quality studies are therefore needed to confirm efficacy and safety and establish an acceptable treatment regimen.

The aim of this proof-of-concept study was to evaluate the contraceptive efficacy, safety, bleeding pattern, and acceptability of once-weekly oral administration of mifepristone 50 mg during 13 menstrual cycles in women aged 18-35.

## METHOD

### Trial design and participants

We performed a prospective, open-label, single arm, multicenter trial in Moldova on 50 mg mifepristone once a week as a contraceptive pill. Participants were recruited from 10 general women’s health clinics in Moldova. The overall objective was to evaluate the contraceptive efficacy and safety of the regimen. The study design was based on the Declaration of Helsinki, ICH E6 (R2) Good Clinical Practice guidelines and European Medicines Agency guidelines.^21^

The study protocol was approved by the Moldovan Medicine and Medical Devices Agency (MMDA) on 24-05-2022 (Rg02-002274), the National Ethics Committee (NEC) on 14-04-2022 and the Swedish Ethical Review Authority (Dnr 2024-08712-01) on 27-01-2025. Amendment 1 was approved by MMDA on 17-06-2022 and NEC on 26-04-2023, amendment 2 by MMDA on 24-05-2024 and NEC on 24-04-2024, and amendment 3 by MMDA on 30-12-2024 and NEC on 29-01-2025. The trial was prospectively registered in the EU Clinical Trials Register (EudraCT 2020-002355-38).

Sexually active women aged 18-35, speaking English, Romanian or Russian, willing to use mifepristone as the only method of contraception for 12 months, with a menstrual cycle of 21-35 days and BMI <35 were eligible for inclusion. Detailed inclusion and exclusion criteria are outlined in Supporting Information Table S1. Written informed consent was obtained before trial entry.

### Study drug intervention

Obninsk (Russia) manufactured the study medication, which was provided in 30-tablet blister packs. Treatment was initiated on days 1-3 of the menstrual cycle. Participants starting on days 4–5 were instructed to use barrier contraception for seven days. Participants switching from oral hormonal contraception started treatment the day after their last pill. Mifepristone was administered once weekly for up to 13 consecutive 28-day cycles. Missed doses were taken as soon as remembered; if more than one day late, barrier contraception was advised for seven days and an additional pregnancy test three weeks later.

### Trial procedures

Screening procedures included a urine hCG-test, transvaginal ultrasound, blood analyses (ALAT, ASAT, bilirubin, creatinine, hemoglobin, LH, FSH, estradiol, and progesterone), cervical smear and a voluntary endometrial biopsy. Study visits were scheduled at screening (visit 1), and at end of month 3 (visit 2), 6 (visit 3), 9 (visit 4) and 12 (visit 5). Vaginal ultrasound and blood analyses were performed at all visits. If ultrasound showed increased endometrial thickness of >15 mm or irregular cystic appearance, an endometrial biopsy was performed, and the participant was followed until endometrial appearance was normal. A voluntary endometrial biopsy was offered at visit 5. Biopsies with suspected pathological changes or progesterone receptor modulator associated endometrial changes (PAEC)^22^ were evaluated by an expert pathologist. ASAT, ALAT ≥2× the upper limit of normal (ULN), or bilirubin >ULN were monitored until normalization. ECG, weight and blood pressure were checked at visit 1, 3 and 5. If a participant was discontinued early due to premature stop of trial, an end-of-trial-visit corresponding to visit 5 was conducted as soon as possible. Pregnancies occurring during the study were followed until the outcome of the pregnancy was known. Participants received 30 tablets of 50 mg mifepristone at visit 1 and 3. Throughout the trial, they performed monthly urine hCG tests and recorded medication intake, intercourse, backup contraception, bleeding, and adverse events (AEs) daily in an electronic diary.

### Sample size

Assuming a true Pearl Index (PI) of 1 pregnancy per 100 woman-years, a total of 12,337 treatment cycles was estimated to be required in order to achieve a difference between the upper and lower limit of the confidence interval (CI) of less than one ^23^. To achieve this number of cycles, we planned to enrol 949 participants, each followed for 13 cycles.

### Primary outcomes

#### Contraceptive efficacy

The primary contraceptive efficacy outcome were perfect-use PIs, expressed as pregnancies per 100 woman-years of exposure, assuming 13, 28-day cycles per year. Typical-use PI was calculated as a secondary outcome. The perfect-use PI included method-failure pregnancies and perfect-use cycles, whereas the typical-use PI included all pregnancies during treatment and perfect, typical, and no-use cycles, including conception cycles. Both PIs were calculated using at-risk cycles (no hormonal contraception and documented unprotected intercourse) and modified at-risk cycles (no hormonal contraception, irrespective of documented intercourse).

Perfect-use required four documented mifepristone doses per cycle taken within a 5–9-day interval. Cycles with an incorrect number of doses or dosing intervals outside this range were classified as typical use. Cycles with explicit evidence of no mifepristone intake were classified as no use, and cycles with insufficient diary information to assess use were classified as non-evaluable.

#### Safety

Safety outcomes included the proportion of women with endometrial thickness >15 mm and the proportion in whom this finding resolved; the proportion experiencing AEs or serious AEs, including symptoms reported in the electronic diary, and severity of AEs; and the proportion with ALAT or ASAT levels ≥3 times the ULN, or bilirubin levels ≥2 times the ULN.

### Secondary outcomes

Secondary outcomes included bleeding patterns (bleeding and spotting days per cycle and proportions of cycles with no bleeding or spotting and spotting only), typical-use PI, PAEC occurrence and resolution, changes in body weight and hemoglobin, occurrence of postmenopausal estradiol levels (<80 pmol/L), and contraceptive acceptability. Spotting was defined as blood loss not requiring sanitary protection. At each follow-up visit, participants rated satisfaction with menstrual bleeding, menstrual pain and the contraception method on a five-point Likert scale ranging from very satisfied to very unsatisfied. They were asked to choose the best and worst aspects of treatment using multiple-choice questions. Acceptability was based on each participant’s last recorded responses.

### Statistical analyses

The database was locked for analysis in March 2026. Statistical analyses were performed using Stata, version 19 (StataCorp, USA), and SPSS, version 31 (IBM, USA).

### Baseline

For baseline characteristics, continuous variables were summarized using means and standard deviations, and categorical variables using frequencies and percentages.

### Analyses of primary outcome and safety

The PI analysis included participants who initiated treatment and documented at least one mifepristone intake in the electronic diary. The treatment period was divided into consecutive 28-day cycles for up to 13 cycles or until discontinuation. Incomplete final cycles were excluded unless conception occurred during the cycle, in which case the cycle was retained and censored at the estimated conception date.

Diary-recorded mifepristone intake and unprotected intercourse were assigned to each cycle. Pregnancies were assigned according to the estimated conception date, determined primarily by ultrasound dating or, when unavailable, from the last menstrual period and reported timing of unprotected intercourse. Conception cycles were classified according to mifepristone use up to conception, subsequent cycles were excluded, and non-evaluable cycles were excluded from PI analyses. PIs were calculated as qualifying pregnancies per 100 woman-years (pregnancies/qualifying cycles × 1300), with two-sided 95% exact Poisson CIs.

Safety analyses used the PI population, with endpoint-specific eligibility criteria. Endometrial and liver safety analyses included participants with at least one post-baseline ultrasound or blood sample, respectively, and outcomes were reported as proportions. AEs were defined as any undesirable event occurring during the study, irrespective of treatment causality. AEs were summarized as the proportion of participants reporting a specific AE ≥10 times and were classified as mild if they did not interfere with daily activities.

### Analyses of secondary outcomes

Bleeding analyses also used the PI population and included perfect- and typical-use cycles with evaluable bleeding data. Mean bleeding and spotting days, and proportions of cycles with no bleeding or spotting and spotting only, were calculated. Baseline and cycle 1 bleeding days were compared using a paired t-test, and bleeding outcomes were visualised using stacked bar charts. Typical-use PIs were calculated according to the described method for PI analysis. Frequencies and proportions for the occurrence and resolution of PAEC, postmenopausal estradiol levels, and contraceptive acceptability were calculated. Changes in hemoglobin and BMI were summarized using means and standard deviations. BMI at the end of treatment was compared with baseline using a paired t-test.

## RESULTS

### Trial population and baseline data

The study was initiated at 10 sites in Moldova. Two sites were closed due to lack of recruitment and one was excluded, in agreement with the institutional review board, due to serious misconduct involving fraudulent data, resulting in seven participating sites contributing to the trial.

Between August 2023 and May 2025, 184 women were screened and 170 were enrolled (Figure 1). Following enrolment, 23 discontinued before treatment initiation, resulting in 147 women starting the trial. Of these, 65 (44%) completed the trial and 82 (56%) discontinued prematurely. One participant had no documented treatment intake and was excluded, resulting in a final analysis and safety population of 146 women. Most discontinuations after treatment initiation (n = 50) resulted from premature trial termination following discontinuation of mifepristone production and misconduct at one study site. In addition, six women were lost to follow-up and 24 women voluntarily withdrew for reasons outlined in figure 1.

**Fig 1.**
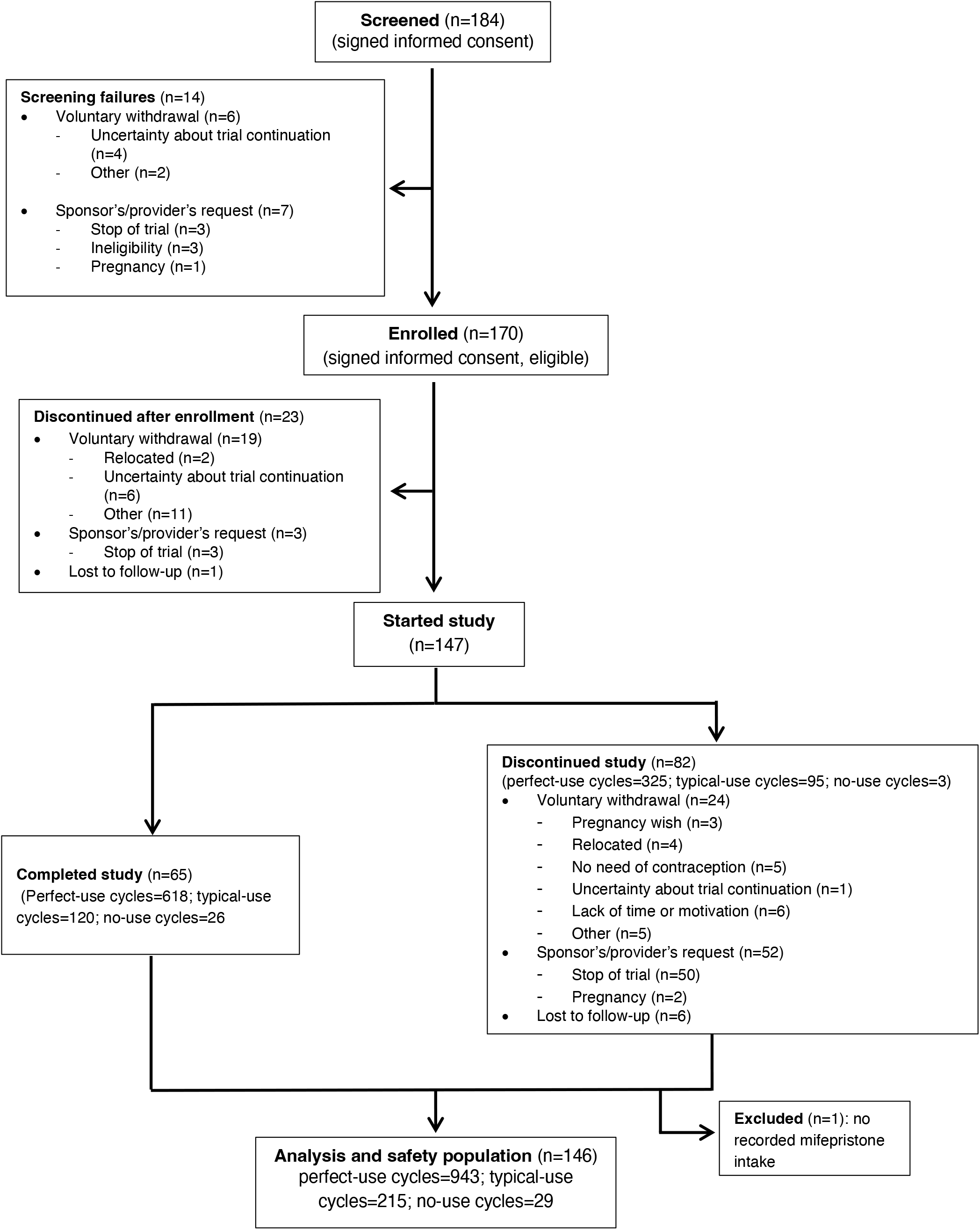
Trial flow-diagram.

Baseline characteristics are presented in Table 1. Mean age was 26.6 years (SD, 4.7) and most participants were nulligravid. The mean menstrual length at baseline was 4.8 days (SD, 1.1). Overall, 24.7% switched from another contraceptive method (32 non-hormonal and four oral hormonal), while approximately 70% had no previous contraceptive use.

**Table 1.** Baseline characteristics of the safety and analysis population. Data are presented as n/N (%) or mean (SD).

| Characteristic | Participants N=146 |
| --- | --- |
| **Age (years), mean (SD)** | 26.6 (4.7) |
| 18-25, n/N (%) | 62/146 (42.5) |
| 26-35, n/N (%) | 84/146 (57.5) |
| **BMI (kg/m<sup>2</sup>), mean (SD)** | 23.1 (3.6) |
| BMI <30, n/N (%) | 136/146 (93.2) |
| BMI 30-35, n/N (%) | 10/146 (6.8) |
| Blood pressure <140/90, n/N (%) | 146/146 (100.0) |
| **Pregnancy history** |  |
| Nulligravid, n/N (%) | 88/146 (60.3) |
| Nulliparous, n/N (%) | 92/146 (63.0) |
| Previous miscarriage, n/N (%) | 6/146 (4.1) |
| Menstrual length (days), mean (SD) | 4.8 (1.1) |
| **Highest level of education** |  |
| No high-school diploma, n/N (%) | 6/146 (4.1) |
| High-school diploma or equivalent, n/N (%) | 16/146 (11.0) |
| University, n/N (%) | 116/146 (79.5) |
| Postgraduate, n/N (%) | 8/146 (5.5) |
| **Smoking status** |  |
| Current smoker, n/N (%) | 13/146 (8.9) |
| **Past contraceptive history** |  |
| Switchers<sup>a</sup>, n/N (%) | 36/146 (24.7) |
| Starters<sup>b</sup>, n/N (%) | 110/146 (75.3) |
| Naïve users<sup>c</sup>, n/N (%) | 102/146 (69.9) |
| **Baseline contraceptive method** |  |
| Oral hormonal contraception, n/N (%) | 4/146 (2.7) |
| Condom, n/N (%) | 0/146 (0.0) |
| Implant, n/N (%) | 0/146 (0.0) |
| Hormonal/copper IUD, n/N (%) | 0/146 (0.0) |
| Vaginal ring, n/N (%) | 0/146 (0.0) |
| Other non-hormonal, n/N (%) | 32/146 (21.9) |
| None, n/N (%) | 110/146 (75.3) |

<sup>a</sup>Switched directly to mifepristone from another contraceptive method.
<sup>b</sup>No baseline contraceptive use, including naïve users with no previous contraceptive use at all.
<sup>c</sup>No previous contraceptive use.

### Contraceptive efficacy

Of the 1,187 evaluable cycles, 943 were classified as perfect use, 215 as typical use, and 29 as non-use (Supporting Information Table S2). Overall, 78% of evaluable cycles were classified as at-risk cycles. An additional 144 cycles were classified as non-evaluable and were consequently excluded from the efficacy analysis.

Four pregnancies occurred during the study (Supporting Information Table S3), of which two occurred during study treatment. The first was an intrauterine pregnancy detected at 4+6 weeks by ultrasound and terminated by medical abortion. Conception occurred during a typical-use cycle with one missed dose and was classified as a typical-use pregnancy. The second was an ectopic pregnancy detected at 7+0 weeks based on estimated conception date, occurred during a perfect-use cycle and was classified as a method failure. The participant underwent laparoscopic salpingectomy. The remaining two pregnancies occurred outside treatment, one during screening and one after treatment completion.

The contraceptive efficacy analysis showed a perfect-use PI of 1.55 (95% CI, 0.04–8.64) for at-risk cycles and 1.38 (95% CI, 0.03–7.68) for modified at-risk cycles (Table 2). The typical-use PI was 2.52 (95% CI, 0.30–9.09) for at-risk cycles and 2.19 (95% CI, 0.27–7.91) for modified at-risk cycles (Table 2).

**Table 2.** Pearl Indices for contraceptive efficacy. Perfect-use and typical-use PI were calculated for both at-risk and modified at-risk cycles. The typical-use PI included perfect-use, typical-use, and no-use cycles, whereas the perfect-use PI included perfect-use cycles only. Exact Poisson 95% CI were calculated for each estimate.

|  | Pregnancies | Cycles | Pearl Index | Exact 95% CI |
| --- | --- | --- | --- | --- |
| Perfect use (at-risk) | 1 | 838 | 1.55 | 0.04–8.64 |
| Perfect use (modified at-risk) | 1 | 943 | 1.38 | 0.03–7.68 |
| Typical use (at-risk) | 2 | 1033 | 2.52 | 0.30–9.09 |
| Typical use (modified at-risk) | 2 | 1187 | 2.19 | 0.27–7.91 |

### Safety

The electronic diary was initiated by 146 participants. AEs recorded in the daily diary a minimum of ten times by at least one participant are presented in Table 3. No AE was reported on at least ten occasions by more than 5% of the participants. The most frequently reported AEs were breast tenderness, mood swings, hot flashes and headache. These AEs were each reported on at least ten occasions by seven of the 146 participants (4.8%). Out of all reported AEs, 91.2% were mild.

**Table 3.** Adverse events reported in the daily diary. Only side effects recorded ≥10 times in the diary per participant are shown in the table.

| Participants reporting a side effect $\geq 10$ times, n/N (%) | Participants N=146 |
| --- | --- |
| Breast tenderness, n/N (%) | 7/146 (4.8) |
| Mood Swings, n/N (%) | 7/146 (4.8) |
| Nausea, n/N (%) | 2/146 (1.4) |
| Hot flashes, n/N (%) | 7/146 (4.8) |
| Headache, n/N (%) | 7/146 (4.8) |
| Dry eyes, n/N (%) | 0/146 (0.0) |
| Other, n/N (%) | 3/146 (2.1) |

Two serious AEs were reported (2/146, 1.4%). As mentioned above, one woman had a laparoscopic salpingectomy due to an ectopic pregnancy. The second serious AE occurred in the participant with a typical-use pregnancy, who was hospitalized for an acute pyelonephritis a few weeks after her medical abortion. This event was assessed as unrelated to mifepristone.

Transvaginal ultrasound examination was performed in 123 study participants during treatment. Eleven participants (11/123, 8.9%) had increased endometrial thickness of > 15 mm on transvaginal ultrasound on at least one occasion. Eight of these women agreed to undergo an endometrial biopsy, which showed PAEC in seven and normal endometrium in one. Among the three participants who declined endometrial biopsy, follow-up ultrasound showed normalization of endometrial thickness in two, whereas the third participant declined further ultrasound examination. Six participants showed PAEC on histological evaluation without previous endometrial irregularities or increased endometrial thickness on ultrasound. No hyperplasia or cellular atypia was detected in any of the endometrial biopsies. One participant with PAEC also had an endometrial polyp without atypia. All 13 participants (13/123, 10.6%) where histology showed PAEC had normal endometrial appearance on ultrasound at follow up.

Regarding liver function values, 133 participants provided blood samples during treatment. Out of these, one participant (0.8%) had elevated levels of liver enzymes and bilirubin elevated >2x the ULN at the 12-month follow-up. The transaminases normalized at follow up and bilirubin decreased to <2x the ULN.

### Bleeding pattern

At baseline, the mean number of bleeding days per cycle was 4.8 (SD, 1.1). This decreased to 1.4 (SD, 1.9) days in the first treatment cycle, corresponding to a 71% reduction in bleeding days from baseline (mean reduction, 3.4 days; 95% CI, 3.0–3.7; *p* <0.001), and remained at a similar level throughout the 13 treatment cycles (Figure 2a and Supporting Information Table S4). Across all treatment cycles, the mean number of bleeding days per cycle was 1.0 (SD, 1.9), while the mean number of spotting days per cycle was 0.7 (SD, 1.4). The proportion of cycles with no bleeding or spotting increased from 45% in treatment cycle 1 to 60% by treatment cycle 13 (Figure 2b). In treatment cycle 13, 85% of cycles involved either no bleeding and no spotting, or spotting only (Figure 2b). Mean hemoglobin levels were 13.2 g/dL (SD, 0.9) at baseline and 13.1 (SD, 0.9) during treatment.

**Fig 2.**
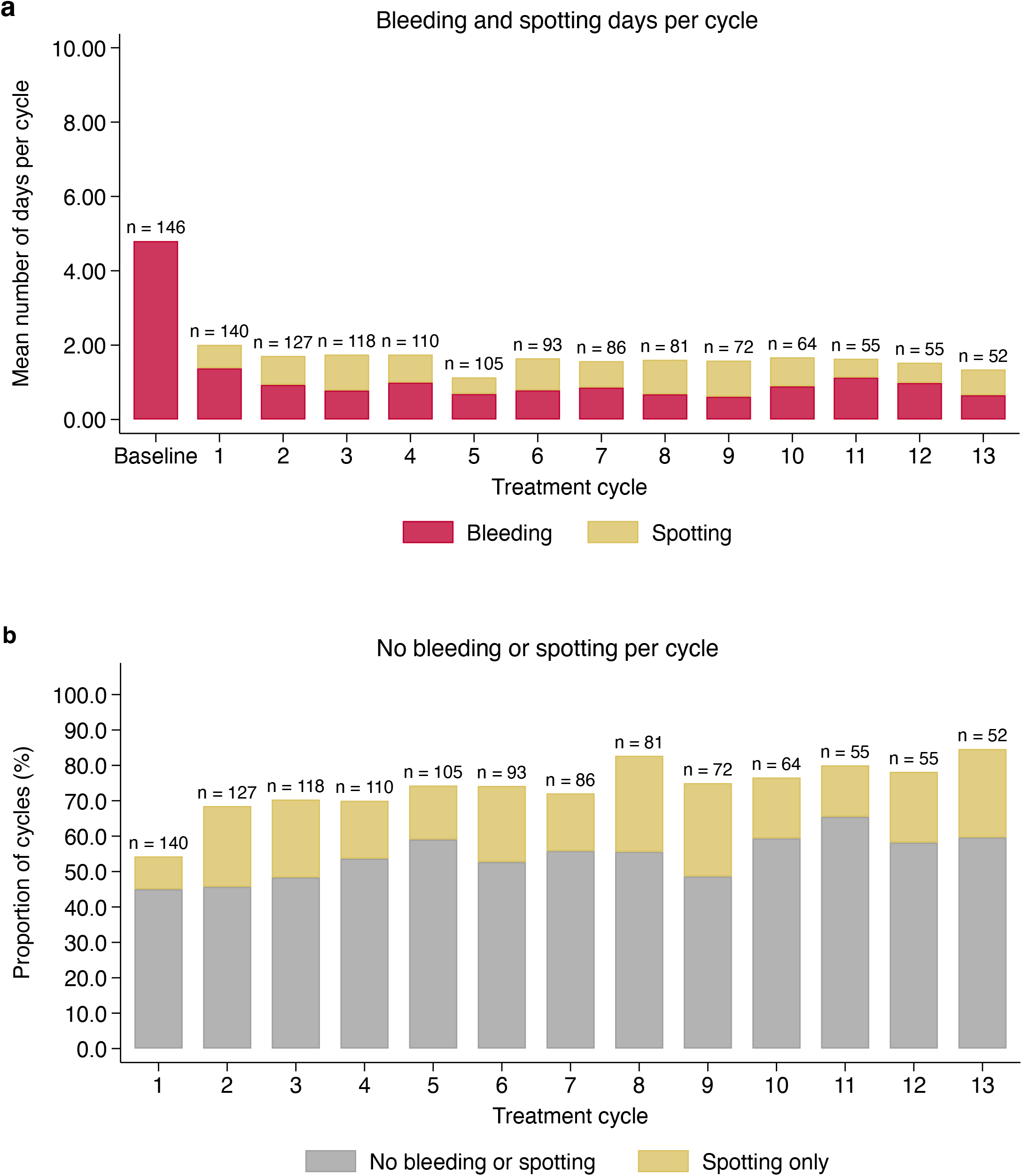
Bleeding pattern during mifepristone treatment. Stacked bar charts showing (**a**) the mean number of bleeding (red) and spotting days (yellow) at baseline and during each treatment cycle and (**b**) the proportion of cycles with no bleeding or spotting (grey) and spotting only (yellow) during each treatment cycle with 50 mg mifepristone administered once weekly. The number above each bar indicates the number of (a) participants or (b) cycles contributing to the respective measurement. Only perfect-use and typical-use cycles were included in both graphs.

### Hormonal levels and weight

Out of the 130 women that evaluated estradiol levels at least once during treatment, no cases of postmenopausal levels of estradiol (<80 picomole/L) were detected. Among women with BMI registered after baseline (n=135), mean BMI decreased from 23.3 (SD, 3.6) at baseline to 22.9 (SD, 3.7) at the last follow-up (95% CI, 0.1–0.6; *p*=0.004).

### Acceptability

Overall, 95.6% (129/135) were satisfied or very satisfied with the contraceptive method, and no participants were unsatisfied (Figure 3). Similar satisfaction rates were seen for menstrual bleeding (123/135, 91.1%) and for menstrual pain (126/135, 93.3%). The best and worst aspects of treatment as reported on the last follow-up visit are summarized in Supporting Information Table S5. Among participants who attended at least one follow-up visit, the most frequently reported best aspect of the treatment was reduced irregular bleeding, reported by 47.8% (65/136). Most participants selected “none” as the worst aspect of treatment (124/136, 91.2%).

**Fig 3.**
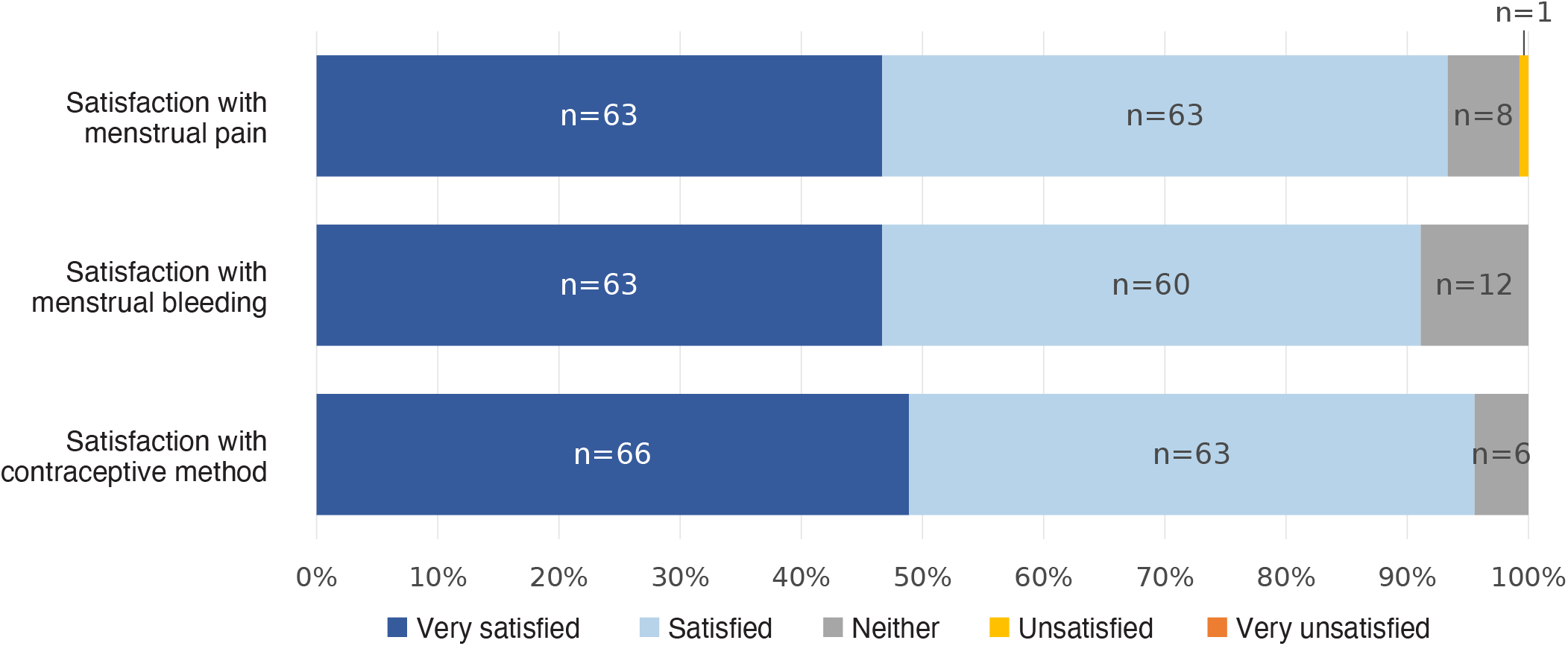
Acceptability. Stacked bar chart showing the proportion of women that were very satisfied (dark blue), satisfied (light blue), neither satisfied or unsatisfied (grey), unsatisfied (yellow) and very unsatisfied (orange) with their menstrual pain, menstrual bleeding and with the contraceptive method. The number in each block represents the number of participants. This figure presents the last recorded response for each participant. Proportions are calculated among participants with available data (N=135).

## DISCUSSION

This study on once-weekly administration of 50 mg mifepristone suggests potential contraceptive efficacy of this treatment regimen. However, the study did not achieve the planned sample size, resulting in a limited number of exposed cycles. The subsequent wide CIs around the estimated PI did not meet the prespecified efficacy criterion. Nonetheless, the study indicates an advantageous safety and bleeding profile, as well as high acceptability among users.

Previous studies have demonstrated contraceptive potential with daily, weekly, and monthly administration of mifepristone and have shown efficacy to vary with dosage regimen.^16,17,19,20,24^ Among weekly regimens, 5 mg once weekly resulted in three pregnancies during 60 treatment cycles.^16^ Similarly, a subsequent study of 5 and 10 mg weekly reported four pregnancies over 64 cycles and three pregnancies during 68 cycles respectively,^17^ indicating insufficient efficacy at these low doses. In contrast, no pregnancies occurred with 25 or 50 mg administered once weekly during a total of 456 cycles.^20^ The present study adds to this previous body of evidence on different contraceptive regimens with mifepristone and, to our knowledge, provides the largest reported exposure of weekly administration to date, comprising 1187 woman-cycles, during which two pregnancies occurred.

The safety findings were generally reassuring, with most AEs being mild and recurrent side effects rare. This is consistent with previous studies of both weekly^16,20^ and daily^19^ mifepristone regimens. Pei et al.^20^ similarly reported uncommon and mild side effects with weekly doses of 25 and 50 mg, including both breast distention and nausea as in the present study. Meanwhile, Marions et al.^16^ reported no disturbing side effects with a 5 mg weekly regimen. Importantly, although the present study observed increased endometrial thickness and PAEC, no endometrial hyperplasia or atypia were identified. PAEC are considered benign, non-physiological changes to the endometrium associated with treatment with progesterone receptor modulators, such as mifepristone, and are reversible after cessation of treatment.^25^ All cases of PAEC and all evaluable cases of increased endometrial thickness normalized at follow-up. These findings are in line with previous studies on the effects on the endometrium following treatment with selective progesterone receptor modulators.^19,25,26^

The two serious AEs comprised one woman having a laparoscopic salpingectomy due to a method failure ectopic pregnancy and an episode of acute pyelonephritis post treatment. No unexpected pattern of serious AEs was observed. However, the limited sample size and evaluable exposure cycles limits conclusions regarding rare or long-term AEs.

Ectopic pregnancies have previously been reported once following the use of 10 mg mifepristone once-a-week.^17^ Daily administration of mifepristone in doses above 5 mg has been shown to inhibit ovulation.^8,19^ Hence, daily administration could be more effective in preventing ectopic pregnancies and should be further evaluated in future studies.

In the present study, mean bleeding days declined from 4.8 at baseline to 1.0 during treatment, similar to Pei et al., who reported a reduction from 5.6 to approximately one day during the final three cycles with weekly 50 mg mifepristone.^20^ In contrast to the previous study, this change occurred already during the first cycle and was sustained throughout treatment. The proportion of cycles with no bleeding or spotting, or spotting only, progressively increased, while spotting days remained stable, indicating an overall shift towards lighter bleeding. Although fewer participants contributed with data in later cycles, the consistent reduction across cycles supports an effect on menstrual bleeding.

The favorable bleeding profile was reflected in the high acceptability of the regimen, with >90% of participants satisfied with the method, menstrual bleeding and pain. Notably, no participant was unsatisfied with the contraceptive method. Reduced irregular bleeding was the most frequently reported benefit of treatment followed by reduced mood changes, breast tenderness and weight loss. These findings indicate that the low-bleeding profile of weekly 50 mg mifepristone may contribute to user satisfaction. More than 90% of participants selected “none” as the worst aspect of the treatment, suggesting few perceived disadvantages. Previous studies of mifepristone for regular contraception have provided limited quantitative data on acceptability.^10,15,16,18–20,27^

This study had several limitations. Recruitment was discontinued before the planned sample size of 949 participants was reached. In addition, only 44% of participants completed the planned 13 treatment cycles. This was primarily due to premature trial discontinuation following the investigational medicinal product becoming unavailable, as well as serious misconduct at one study center. Consequently, there was insufficient precision to meet the prespecified efficacy criterion of a difference between the upper and lower CI of <1. The variable duration of exposure to mifepristone also limits the assessment of longer-term safety, bleeding pattern and acceptability. Moreover, the study did not include a comparator contraceptive method. Hence, comparison of contraceptive efficacy, safety profile and acceptability in relation to existing contraceptive methods was not possible.

## CONCLUSION

We conclude that weekly administration of 50 mg mifepristone showed potential contraceptive efficacy, favorable safety and bleeding profiles, and high acceptability. However, the limited number of treatment cycles resulted in an imprecise estimate of the PI, limiting certainty regarding contraceptive efficacy and warranting further evaluation in larger studies.

## Ethics statement

The study protocol was approved by the Moldovan Medicine and Medical Devices Agency (MMDA) on 24-05-2022 (document number Rg02-002274), the National Ethics Committee (NEC) on 14-04-2022, and the Swedish Ethical Review Authority (Dnr 2024-08712-01) on 27-01-2025. Amendment 1 was approved by MMDA on 17-06-2022 and NEC on 26-04-2023, amendment 2 by MMDA on 24-05-2024 and by NEC on 24-04-2024, and amendment 3 by MMDA on 30-12-2024 and by NEC on 29-01-2025. The trial was prospectively registered in the EU Clinical Trials Register on 02-02-2021 (EudraCT Number: 2020-002355-38, URL: https://www.clinicaltrialsregister.eu/ctr-search/search?query=mife50). Participant enrollment was initiated on 07-08-2023. Written informed consent was obtained before enrollment.

## Acknowledgements

We would like to thank Professor Alistair Williams, Professor of Gynecological Pathology at the Queens Medical Research Institute, University of Edinburgh, for his contribution as consulting pathologist and assessments of PAEC, and national coordinating investigator Dr Rodica Commendant, the local investigators, and other collaborators.

## Author contribution

Kristina Gemzell Danielsson contributed to the study conception and design, protocol development, and study funding. Caroline Frisendahl contributed to the protocol development, data quality controls, and data cleaning. Agnes Herré and Caroline Frisendahl contributed to planning and conducting the statistical analysis, interpretation of the results, and drafting the manuscript. Agnes Herre and Caroline Frisendahl directly accessed and verified the individual participant-level data reported in the manuscript, and all authors had access to the data. Karin Brandell and Kristina Gemzell Danielsson contributed to planning the statistical analysis and interpretation of the findings, revised the manuscript, and approved the final version.

## Data Availability statement

De-identified data may be available after publication upon reasonable request to the corresponding author.

## Abbreviations

AE(s): Adverse event(s)
CI: Confidence Interval
MMDA: Medicine and Medical Devices Agency
PAEC: Progesterone receptor modulator associated changes
PI: Pearl Index
ULN: Upper limit of normal

## Legends of supporting information

**Table S1. Inclusion and exclusion criteria for the trial participants.**

**Table S2. Cycle use and risk classification.** The 28-day cycles were classified as perfect use, typical use, no use, or non-evaluable based on mifepristone intake. Cycles were further classified as at risk if unprotected sexual intercourse occurred during the cycle.

**Table S3. Characteristics of pregnancies occurring during the study period.**

**Table S4. Bleeding and spotting days per treatment cycle.** Results are presented as the mean number of bleeding and spotting days per treatment cycle, with standard deviations (SDs) in parentheses.

**Table S5. Best and worst aspects of treatment.** At each follow-up visit, participants were asked to choose the best and worst aspects of treatment using multiple-choice questions, with the option to select more than one response. This table presents the last recorded response for each participant. Percentages were calculated using the total number of participants with a recorded response as denominator (N=136).

## References

1 Ali MM, Cleland J, Shah IH. Causes and consequences of contraceptive discontinuation: evidence from 60 demographic and health surveys. 2012 https://iris.who.int/handle/10665/75429 (accessed April 6, 2026).

2 Fiala C, Danielsson KG. Review of medical abortion using mifepristone in combination with a prostaglandin analogue. Contraception. 2006; 74: 66–86.

3 Gemzell-Danielsson K, Swahn M-L, Svalander P, Bygdeman M. Early luteal phase treatment with mifepristone (RU 486) for fertility regulation. 1993.

4 Spitz IM. Mifepristone: Where do we come from and where are we going? Clinical development over a quarter of a century. Contraception. 2010; 82: 442–52.

5 Llut JH, Garzo G, Morris S, et al. Disruption of Follicular Maturation and Delay of Ovulation after Administration of the Antiprogesterone RU486*. 1987.

6 Luukkainen T, Heikinheimo O, Haukkamaa M, Liihteenmiiki P. FERTILITY AND STERILITY Inhibition of folliculogenesis and ovulation by the anti progesterone RU 486*t. 1988.

7 Batistat MC, Cartledge TP, Zellmer AW, et al. Evidence for a Critical Role of Progesterone in the Regulation of the Midcycle Gonadotropin Surge and Ovulation*. 1992.

8 Ledger WL, Sweeting VM, Hillier H, Baird DT. Inhibition of ovulation by low-dose mifepristone (RU 486). 1992.

9 Gemzell-Danielsson K, Svalander P, Swahn M-L, Johannisson E, Bygdeman M. Effects of a single post-ovulatory dose of RU486 on endometrial maturation in the implantation phase. 1994.

10 Hapangama DK, Brown A, Glasier AF, Baird DT. Feasibility of administering mifepristone as a once a month contraceptive pill. 2001.

11 Bartlett TE, Evans I, Jones A, et al. Antiprogestins reduce epigenetic field cancerization in breast tissue of young healthy women. Genome Med 2022; 14. DOI:10.1186/s13073-022-01063-5.

12 Engman M, Skoog L, Söderqvist G, Gemzell-Danielsson K. The effect of mifepristone on breast cell proliferation in premenopausal women evaluated through fine needle aspiration cytology. Human Reproduction 2008; 23: 2072–9.

13 Utjés D, Alkasalias T, Cameron ST, et al. The potential role of mifepristone in breast cancer prevention: beyond medical abortion. The Lancet Obstetrics, Gynaecology, & Women’s Health 2025; : 100013.

14 Marions L, Viski S, Danielsson KG, et al. Contraceptive efficacy of daily administration of 0.5 mg mifepristone. 1999.

15 Lakha F, Ho PC, Van der Spuy ZM, et al. A novel estrogen-free oral contraceptive pill for women: Multicentre, double-blind, randomized controlled trial of mifepristone and progestogen-only pill (levonorgestrel). Human Reproduction 2007; 22: 2428–36.

16 Marions L, Danielsson KG, Swahn M-L, Bygdeman M. Contraceptive efficacy of low doses of mifepristone. 1998.

17 Cheng J, Weng L, Han X. Study on weekly low doses of mifepristone for contraception. Zhonghua Fu Chan Ke Za Zhi 2001; 36: 424–7.

18 Godfrey EM, Mawson JT, Stanwood NL, Fielding SL, Schaff EA. Low-dose mifepristone for contraception: a weekly versus planned postcoital randomized pilot study. Contraception 2004; 70: 41–6.

19 Brown A, Cheng L, Lin S, Baird DT. Daily Low-Dose Mifepristone Has Contraceptive Potential by Suppressing Ovulation and Menstruation: A Double-Blind Randomized Control Trial of 2 and 5 mg per Day for 120 Days. 2002 https://academic.oup.com/jcem/article/87/1/63/2846605.

20 Pei K, Xiao B, Jing X, Lu S, Wei L, Zhao H. Weekly contraception with mifepristone. Contraception 2007; 75: 40–4.

21 The EMEA CHMP guideline on clinical investigation of steroid contraceptives in women https://www.ema.europa.eu/en/clinical-investigation-steroid-contraceptives-women-scientific-guideline (accessed Aug 29, 2026).

22 Mutter GL, Bergeron C, Deligdisch L, et al. The spectrum of endometrial pathology induced by progesterone receptor modulators. Modern Pathology 2008; 21: 591–8.

23 Gerlinger C, Endrikat J, Van Der Meulen EA, Dieben TOM, Düsterberg B. Recommendation for confidence interval and sample size calculation for the Pearl Index. European Journal of Contraception and Reproductive Health Care 2003; 8: 87– 92.

24 Glasier A, Thong KJ, Dewar M, Mackie M, Baird DT. Mifepristone (RU 486) compared with High-dose Estrogen and Progestogen for Emergency Postcoital Contraception. 1992.

25 Williams ARW, Bergeron C, Barlow DH, Ferenczy A. Endometrial morphology after treatment of uterine fibroids with the selective progesterone receptor modulator, ulipristal acetate. International Journal of Gynecological Pathology 2012; 31: 556–69.

26 Chodankar RR, Murray A, Nicol M, Whitaker LHR, Williams ARW, Critchley HOD. The endometrial response to modulation of ligand-progesterone receptor pathways is reversible. Fertil Steril 2021; 116: 882–95.

27 Agarwal M, Das V, Agarwal A, Pandey A, Srivastava D. Evaluation of mifepristone as a once a month contraceptive pill. Am J Obstet Gynecol 2009; 200. DOI:10.1016/j.ajog.2008.09.005.

