## Supplementary material for "Contraceptive efficacy of 50 mg Mifepristone once-a-week: an open label, single-arm, multicenter clinical trial in Moldova": Table S1

### **Table S1. Inclusion and exclusion criteria for the trial participants.**

| **Inclusion criteria** | - Female - Aged 18-35 - Living in Moldova and speaking either Romanian, English or Russian - Regular menstrual cycles with menstrual cycle length of 21-35 days - After Depo-Provera (3-month injectable) at least 3 cycles of 21-35 days - No desire to become pregnant within the next 12 months - Be willing to use mifepristone as the only contraceptive method during the study - Be willing to use the study drug as the only method of contraception - Expecting to have unprotected intercourse at least once a month with a partner who is not sterilized - BMI < 3 - Be able to participate in the scheduled visits - Be willing to fill in daily diary via app and 6-monthly questionnaires via app and online or paper - Be willing to provide signed and dated informed consent. |
| --- | --- |
| **Exclusion criteria** | - Current pregnancy - Known infertility, subfertility or previous ectopic pregnancy - Current pregnancy - Lactation - Undiagnosed abnormal vaginal bleeding - Abnormal smear at screening which requires colposcopy and/or biopsy according to the Moldova national guideline for abnormal pap-test without colposcopic evaluation - Any previous or current malignancy including breast cancer - Previous or current liver illness, infection or ASAT, ALAT, or bilirubin > 2 ULN - Family history of endometrial cancer - Known BRCA genome mutation - Intracavitary abnormalities on vaginal ultrasound at screening, including intracavitary polyps or myomas, irregular cystic endometrium, endometrium > 15 mm, or an obvious aspect of a hydrosalpinx - Signs of current endometritis, incomplete abortion after miscarriage or induced abortion, retained placental rests after delivery or pelvic inflammatory disease (PID) - History of gastric bypass - Known allergy to mifepristone - Using high doses of corticosteroids or any drugs that may interact with mifepristone - these include hydantoins (e.g. phenytoin), barbiturates (e.g. phenobarbital), primidone, carbamazepine, rifampicin, oxcarbazepine, topiramate, rifabutin, felbamate, ritonavir, nelfinavir, griseofulvin and products containing St. John's wort (Hypericum perforatum); - Treatment with another investigational drug (until next normal period after stopping any other trial) - Unable to comply with the trial protocol - Undiagnosed reason for severe anaemia or increased creatinine - Severe hypertension - 180 mm Hg or more systolic, or/and 110 mm Hg or more diastolic (hypertension with medical treatment allowed). |
