## Supplementary material for "Contraceptive efficacy of 50 mg Mifepristone once-a-week: an open label, single-arm, multicenter clinical trial in Moldova": Table S2

**Table S2. Cycle use and risk classification.** The 28-day cycles were classified as perfect use, typical use, no use, or non-evaluable based on mifepristone intake. Cycles were further classified as at risk if unprotected sexual intercourse occurred during the cycle.

| **Cycle use classification** | **Modified**  **at-risk** | **At-risk** | **Total** |
| --- | --- | --- | --- |
| Perfect use | 105 | 838 | 943 |
| Typical use | 33 | 182 | 215 |
| No use | 16 | 13 | 29 |
| Non-evaluable | 144 | 0 | 144 |
| **Total** | 298 | 1033 | 1331 |
