## Supplementary material for "Contraceptive efficacy of 50 mg Mifepristone once-a-week: an open label, single-arm, multicenter clinical trial in Moldova": Table S3

### **Table S3. Characteristics of pregnancies occurring during the study period.**

| **Classification** | **Conception cycle** | **Location** | **Gestational length (weeks + days)** | **Outcome** |
| --- | --- | --- | --- | --- |
| Method failure | Cycle 2 | EP | 7+0 | Laparoscopic salpingectomy |
| Typical use | Cycle 4 | IUP | 4+6 | Medical abortion |
| Before treatment | ---- | IUP | 6+2 | Surgical abortion |
| After treatment | ---- | IUP | 5+3 | Surgical abortion |
