## Supplementary material for "Contraceptive efficacy of 50 mg Mifepristone once-a-week: an open label, single-arm, multicenter clinical trial in Moldova": Table S4

**Table S4. Bleeding and spotting days per treatment cycle.** Results are presented as the mean number of bleeding and spotting days per treatment cycle, with standard deviations (SDs) in parentheses.

| **Treatment cycle** | **Cycles**  **contributing** | **Mean bleeding days (SD)** | **Mean spotting days (SD)** |
| --- | --- | --- | --- |
| 1 | 140 | 1.38 (1.85) | 0.63 (1.05) |
| 2 | 127 | 0.93 (1.71) | 0.78 (1.28) |
| 3 | 118 | 0.78 (1.50) | 0.97 (1.56) |
| 4 | 110 | 1.00 (1.91) | 0.75 (1.78) |
| 5 | 105 | 0.69 (1.42) | 0.45 (0.93) |
| 6 | 93 | 0.78 (1.56) | 0.86 (1.61) |
| 7 | 86 | 0.86 (1.57) | 0.71 (1.38) |
| 8 | 81 | 0.68 (1.84) | 0.93 (1.66) |
| 9 | 72 | 0.61 (1.27) | 0.97 (1.58) |
| 10 | 64 | 0.89 (1.89) | 0.78 (1.64) |
| 11 | 55 | 1.13 (3.55) | 0.51 (1.14) |
| 12 | 55 | 0.98 (2.53) | 0.55 (1.10) |
| 13 | 52 | 0.65 (2.21) | 0.69 (1.37) |
