## Supplementary material for "Contraceptive efficacy of 50 mg Mifepristone once-a-week: an open label, single-arm, multicenter clinical trial in Moldova": Table S5

**Table S5. Best and worst aspects of treatment.** At each follow-up visit, participants were asked to choose the best and worst aspects of treatment using multiple-choice questions, with the option to select more than one response. This table presents the last recorded response for each participant. Percentages were calculated using the total number of participants with a recorded response as denominator (N=136).

|  | **Participants (N=136)** |
| --- | --- |
| **Best aspects of treatment** |  |
| Reduced irregular bleeding, n/N (%)  Weight loss, n/N (%)  Reduced nausea/vomiting, n/N (%)  Reduced acne, n/N (%)  Reduced breast tenderness, n/N (%)  Reduced mood changes, n/N (%)  None, n/N (%)  Other, n/N (%) | 65/136 (47.8)  37/136 (27.2)  13/136 (9.6)  9/136 (6.6)  39/136 (28.7)  40/136 (29.4)  38/136 (27.9)  13/136 (9.6) |
| **Worst aspects of treatment** |  |
| None, n/N (%)  Irregular bleeding, n/N (%)  Weight gain, n/N (%)  Nausea/vomiting, n/N (%)  Acne, n/N (%)  Breast tenderness, n/N (%)  Mood changes, n/N (%)  Other, n/N (%) | 124/136 (91.2)  4/136 (2.9)  2/136 (1.5)  0/136 (0.0)  0/136 (0.0)  4/136 (2.9)  0/136 (0.0)  1/136 (0.7) |
